# Irreversible electroporation associated with improved overall survival vs standard of care for stage 3 pancreatic ductal adenocarcinoma

**DOI:** 10.64898/2026.05.19.26353144

**Authors:** Robert C. G. Martin, Rebekah Ruth White, Malcolm M. Bilimoria, Govindarajan Narayanan, Michael D. Kluger, David A. Iannitti, Patricio M. Polanco, Chet W. Hammill, Sean P. Cleary, Robert Evans Heithaus, Theodore Welling, Carlos H. F. Chan

## Abstract

**Background:** Emerging evidence suggests irreversible electroporation (IRE) with standard-of-care (SOC) chemotherapy may improve survival in patients with Stage 3 pancreatic ductal adenocarcinoma (PDAC) when compared to SOC alone. This study evaluates the overall survival (OS) and progression-free survival (PFS) of Stage 3 PDAC patients treated with SOC plus IRE with the NanoKnife System versus SOC alone.

**Methods:** This prospective, multicenter study included two cohorts from the DIRECT registry: an IRE cohort from sites offering IRE as part of clinical care, and a comparator SOC cohort of prospectively enrolled and contemporaneous retrospective patients. Enrollment spanned 08/05/2019 to 02/05/2023, with follow-up through at least 24 months, death, or loss to follow-up. Included were 137 patients (99 IRE; 38 SOC), aged ≥18 years with Stage 3 PDAC and no progression after three months of SOC therapy.

**Results:** Median (interquartile range) time from diagnosis to enrollment was 8 (6-10) months for IRE and 4 (3-6) for SOC (p<0.0001). Median OS and PSF from enrollment were 18 (95% confidence interval [CI]: 15-24) months and 9 (95% CI: 7-12) months for IRE, and 10 (95% CI: 8-14) months and 6 (5-8) months for SOC, respectively (p<0.0001 and p=0.009). Adverse events occurred in 80% (79/99) of IRE patients and 95% (36/38) of SOC patients; 29% (29/99) of the IRE cohort experiencing an IRE-related adverse event.

**Conclusions:** IRE was associated with improved OS versus SOC alone and may be an effective consolidative treatment for Stage 3 PDAC after three months of induction chemotherapy.

## Introduction

Pancreatic ductal adenocarcinoma (PDAC) is the most common form of pancreatic cancer, the third leading cause of cancer related death in the United States, with a 5-year survival rate of around 10% [1]. At the time of diagnosis, only 15-20% of pancreatic cancers are considered resectable [2]. Treatment is complex for patients with Stage 3 pancreatic cancer (Stage 3 PC) with systemic chemotherapy with or without radiation as the current standard of care (SOC). Recent evidence suggests 5-fluorouracil, leucovorin, irinotecan, and oxaliplatin (FOLFIRINOX) and combination therapy with gemcitabine and nab-paclitaxel have improved efficacy over gemcitabine alone and are considered the optimal first line therapy [3].

For patients with Stage 3 locally advanced unresectable disease, irreversible electroporation (IRE) is a non-thermal ablation method that was cleared by the United States Food and Drug Administration (FDA) in 2010 under 510(k) K102329 and uses high-voltage short electrical pulses to change the permeability of the cell membrane, leading to cell death [4–6]. Due to its non-thermal mechanism of action, since 2012 IRE has become a potential treatment option for Stage 3 PC as it allows for the ablation of tumors while preserving surrounding vital structures [7,12]. There are two operative clinical uses of IRE for pancreatic cancer: IRE for the ablation of unresectable lesions (in-situ ablation) and margin accentuation when resection is feasible, but there are concerns for a positive margin at vital structures [8]. Historically, unresectable Stage 3 PC had a low median survival of 9-16 months depending on the treatment used [9,10]. Emerging evidence suggests IRE may improve overall survival (OS) in this patient population [11–15]. A large, prospective registry study of 200 patients with locally advanced pancreatic cancer (LAPC) reported a median OS from diagnosis of 25 months [16]. Furthermore, PANFIRE-2, a prospective, phase II trial, studied the safety and efficacy of percutaneous IRE for LAPC and reported a median OS from diagnosis of 17 months which exceeded the target median OS for patients receiving no induction chemotherapy or gemcitabine-based induction chemotherapy (12 months) and for those receiving induction FOLFIRINOX (15 months) [17].

As a United States FDA Investigational Device Exemption (IDE) study, DIRECT is a registry study aimed to evaluate the effectiveness and safety of SOC plus IRE with the NanoKnife System compared to SOC alone for the treatment of Stage 3 PDAC.

A previous safety publication from the DIRECT registry demonstrated that the use of IRE for curative intent tumor ablation in combination with induction chemotherapy has equivalent morbidity and 90-day mortality rates when compared to SOC chemotherapy alone [18]. This paper reports the OS and progression-free survival (PFS) of these cohorts.

## Materials and Methods

### Study Design and Patient Selection

The DIRECT Registry (Clinicaltrials.gov NCT03899649) study is a prospective, observational registry of patients in the United States from 18 clinical sites. This study was not randomized and not blinded. Enrollment began in 08/05/2019 and ended in 02/05/2023, and patients were followed through at least 24 months, death, or loss to follow-up. The study included patients with Stage 3 Pancreatic ductal adenocarcinoma (PDAC) undergoing IRE with the NanoKnife System in addition to SOC in accordance with each participating sites’ guidelines, as well as a comparator cohort of patients with Stage 3 PDAC who received at least three months of SOC but did not receive IRE. Patients treated with IRE were prospectively enrolled from sites that provide IRE as part of clinical care (IRE sites) while the comparator cohort was comprised of prospectively enrolled patients and contemporaneous retrospective data collected on patients at DIRECT sites undergoing SOC. For the retrospective SOC patients, “enrollment” was defined as the point at which patients became eligible for the study after 3 months of SOC therapy (i.e., were re-staged as Stage 3 to confirm they were neither down staged nor progressed following treatment). The SOC cohort was enrolled from 26/09/2019 to 17/06/2024. All enrolled patients (IRE and SOC) finished the study and there was no attrition. The attrition diagram and study design scheme are available in S1 and S2 Fig, respectively.

The site enrollment periods were as follows: University of Louisville (10/04/2021 to 27/09/2023), Northwest Community Healthcare (15/08/2019 to 27/09/2023), University of South Florida and Tampa General Hospital (20/12/2019 to 07/12/2022), Baptist Hospital of Miami and Miami Cancer Institute (22/11/2019) to (27/09/2023), University of California San Diego (17/04/2020 to 27/09/2023), Atrium Health (24/02/2020 to 16/12/2022), Columbia University (09/06/2020 to 27/09/2023), UT Southwestern Medical Center (30/05/2020 to 02/12/2022), Barnes-Jewish Hospital and Washington University School of Medicine (08/01/2020 to 01/12/2022), Mayo Clinic Minnesota (05/03/2021 to 27/09/2023), Mayo Clinic Florida 04/02/2021 to 01/12/2022), WellStar Summit Surgical (30/03/2020 to 02/12/2022), Capital Health System, Inc (12/05/2020 to 01/12/2022), St. Luke’s University Health Network (02/06/2020 to 01/12/2022), University of Iowa (21/10/2020 to 17/06/2024), Augusta University (03/12/2020 to 25/05/2022), University of Florida Gainesville (15/05/2021 to 07/12/2022), and NYU Langone Health (26/03/2021 to 21/08/2023).

Males and Females, 18 years of age or older with pathologically confirmed American Joint Committee on Cancer (AJCC) stage 3 PDAC with Eastern Cooperative Oncology Group (ECOG) performance status of 0 or 1 and no evidence of disease progression based on National Comprehensive Cancer Network (NCCN) guidelines after completing three months of SOC chemotherapy were eligible for inclusion in the study. Exclusion criteria included participation in an interventional trial for pancreatic cancer during the data collection period and inability to tolerate general anesthetic with full skeletal muscle blockade. Complete eligibility criteria are available in S1 Methods. Patients in both cohorts were enrolled at the completion of three months of initial SOC treatment for stage 3 PDAC. Patients continued to receive SOC chemotherapy after completing the required three months of treatment as per the treating principal investigator and/or treating physician.

### Study Procedure

Patients in the IRE cohort were treated with IRE using the NanoKnife System (AngioDynamics, Inc., Latham, NY, USA). IRE protocol (Clinicaltrials.gov NCT03899649) and patient selection with the NanoKnife System has been previously described [18,19]. The Prior to study initiation, all participating physicians at IRE sites were trained on optimal tissue specific use procedures for the NanoKnife System, including procedure settings and post-procedure follow-up. Sites were requested to adhere to guidelines for the treatment and monitoring of patients, testing markers (Carbohydrate Antigen 19-9 [CA 19-9]) and performing imaging (computed tomography [CT] or magnetic resonance imaging [MRI]) at least every three to four months during the first two years [20].

The study received approval from the Western Institutional Review Board (IRB approval 2019-0965) and local IRBs. All prospective patients provided written informed consent. The retrospective cohort was also prospectively consented for data collection. The study was conducted in accordance with the International Conference on Harmonization’s Good Clinical Practice guidelines.

### Assessments and Data Collection

Data elements were entered into electronic case report forms at baseline, at the index hospitalization for IRE patients, and at 3-month intervals until study exit (death, withdrawal, loss to follow-up) or a minimum of 24 months of follow up was obtained. At each evaluation from the time of enrollment, sites were instructed to determine whether an adverse event (AE) occurred including relationship to underlying disease, procedure, and/or treatments. AEs were graded based on Common Terminology Criteria for Adverse Events (CTCAE) grade 1 (mild) to 5 (death).

### Endpoints

The primary effectiveness endpoint was OS defined as time in months from enrollment to date of death for any reason. Patients alive or lost to follow-up at the time of analysis were censored at their last date of follow-up.

Secondary effectiveness endpoints included PFS defined as the time in months from enrollment to the date of first observed disease progression or death from any cause, if death occurs without documented disease progression, as well as OS defined as time in months from diagnosis to date of death for any reason.

An independent Clinical Endpoint Adjudication Committee (CEC) was established during this study. The main purpose was to review and adjudicate individual patient AEs and serious AEs (SAEs). The CEC consisted of three members with expertise in pancreatic cancer management. None of the members had direct involvement in the conduct of the study or had any conflicts of interest. A Data Monitoring Committee was also established to review cumulative study data to evaluate safety, study conduct, and scientific validity and integrity of the trial. The Data Monitoring Committee (DMC) consisted of three physicians and a statistician who were independent from the trial and had no conflicts of interest.

AEs are presented including relationship to treatments received and AEs CTCAE grade 3 or higher.

### Statistical Analysis

Statistical analysis was performed using Statistical Analysis System (RRID:SCR_008567). Data for continuous variables are presented using descriptive statistics including the mean, standard deviation (SD), median, and interquartile range (IQR). Categorical variables are presented using descriptive statistics including frequencies and percents. IRE cohort analyses were stratified by procedure type (open vs. percutaneous). In this study, missing data were not imputed nor handled with last observation carried forward. In general, data were analyzed as they were recorded.

Analyses comparing OS time between IRE and SOC cohorts were based on Log-rank test. Kaplan-Meier plots were used to show the distribution of the events, with loss to follow-up censored at the time of last contact. Median time in months and the associated 95% confidence intervals (CIs) are presented for both IRE and SOC cohorts. As a sensitivity analysis to the stratified log-rank test, the Cox proportional hazard regression model was applied.

### Role of the Sponsor

The study sponsor (AngioDynamics, Inc.) provided financial and logistical support but had no role in data collection or data interpretation. The academic investigators had full access to the complete dataset and retained final responsibility for data interpretation and the decision to submit the manuscript for publication.

## Results

### Patient Characteristics and Procedure Details

A total of 137 patients were included in the analysis, with 99 in the IRE cohort and 38 in the SOC cohort. Within the IRE cohort, 87 patients received open IRE and 12 received percutaneous IRE. The median (IQR) duration of follow-up was 16 (8-27) months for the IRE cohort and 10 (7-16) months for the SOC cohort. The median (IQR) age was 65 (58-71) years in the IRE cohort and 67 (61-75) years in the SOC cohort. Median (IQR) time from diagnosis to enrollment was 8 (6-10) months for the IRE cohort compared with 4 (3-6) months in the SOC cohort (p<0.0001). Regarding induction (first line) chemotherapy, most patients in both cohorts received FOLFIRINOX; 51% (50/99) in the IRE arm and 58% (22/38) in the SOC arm, with a median (IQR) of 3 (1-8) cycles in the IRE cohort and 4 (4-7) cycles in the SOC cohort (p=0.79). Patients in the IRE cohort were more likely to have had prior radiation therapy: 38% (38/99) for IRE vs. 3% (1/38) for SOC, p<0.0001. Patient and disease characteristics are presented in Table 1 and were similar between cohorts. The procedure details for the IRE cohort are presented in Table 2.

**Table 1.**
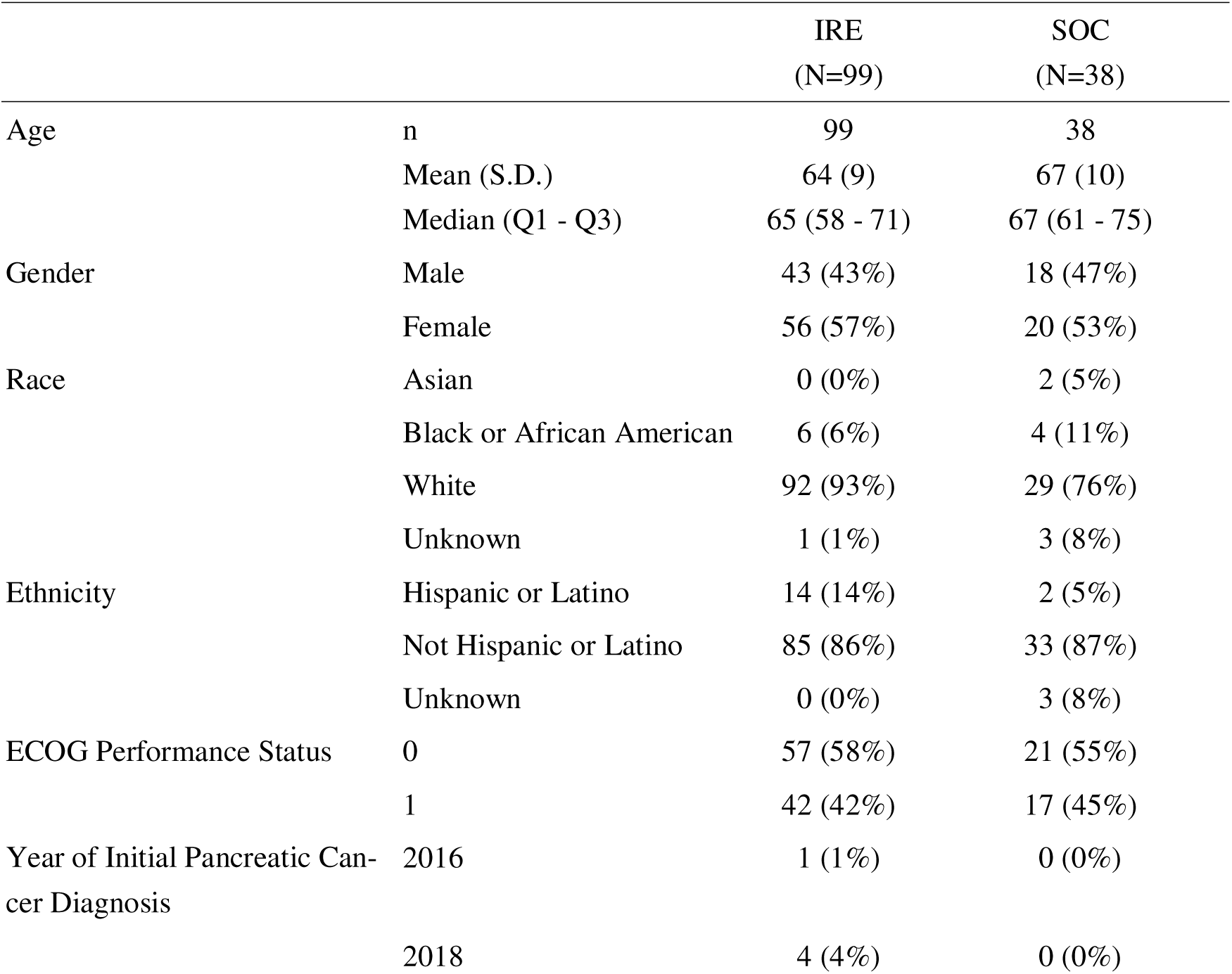
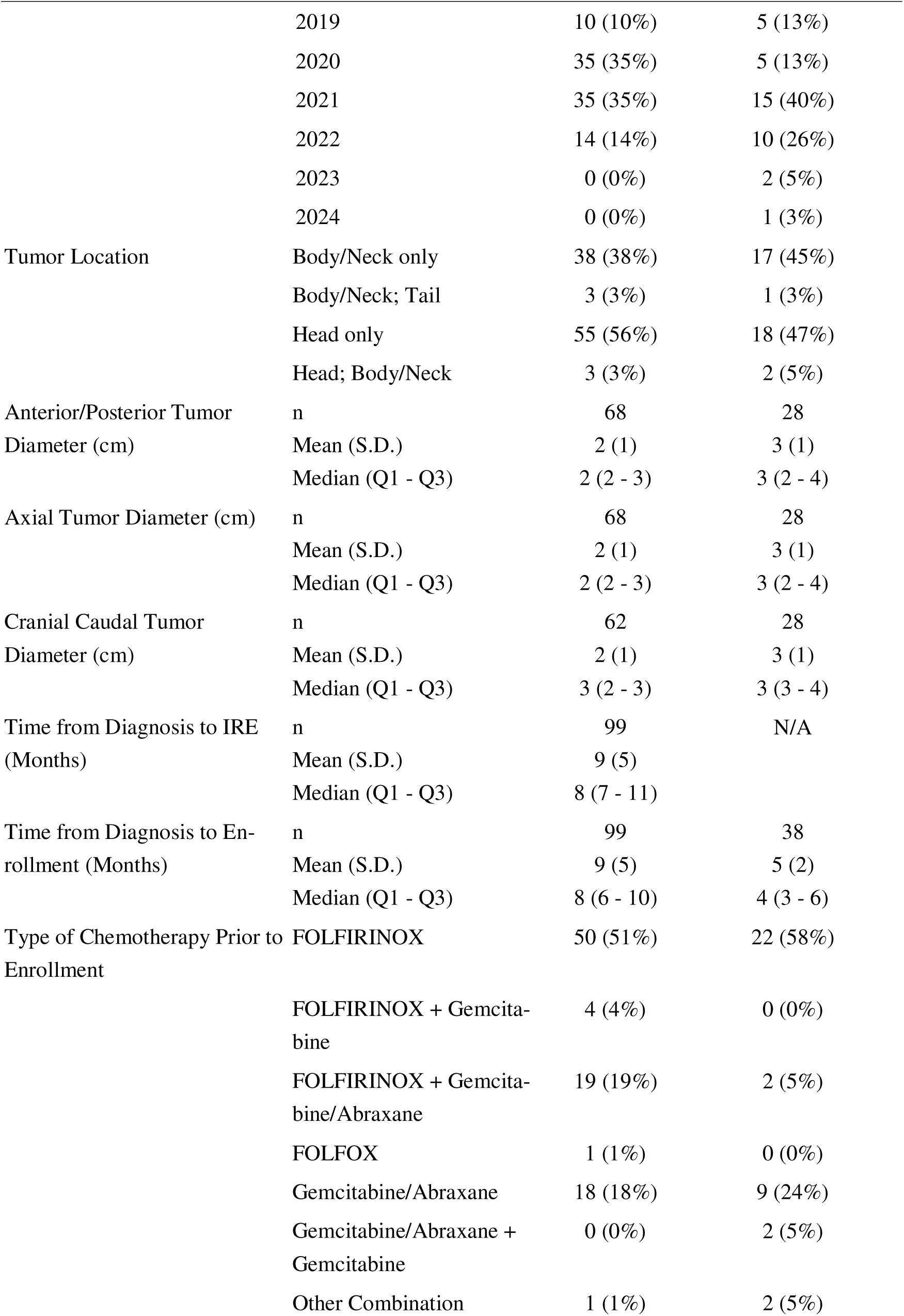
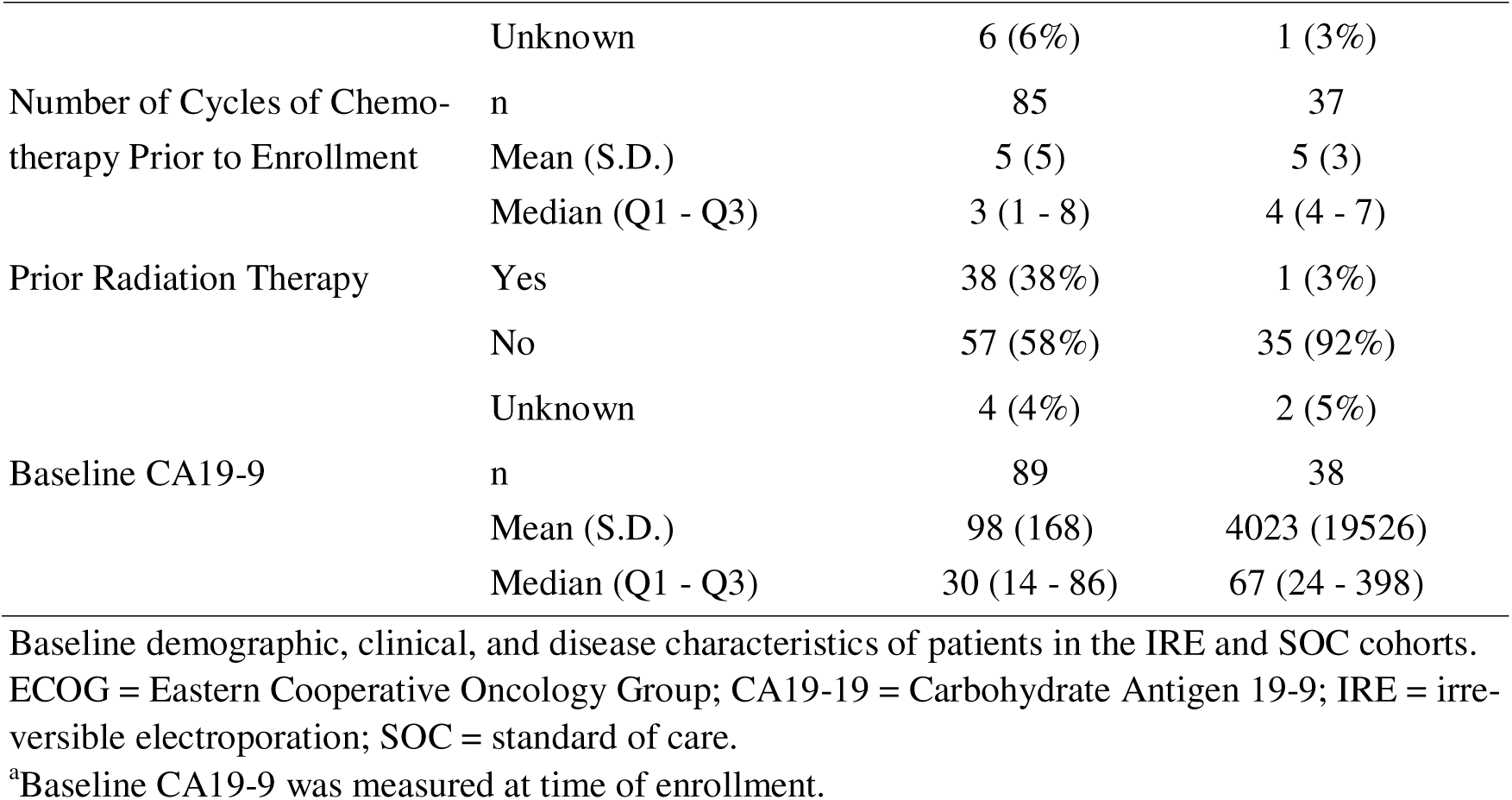
Patient and disease characteristics.

**Table 2.**
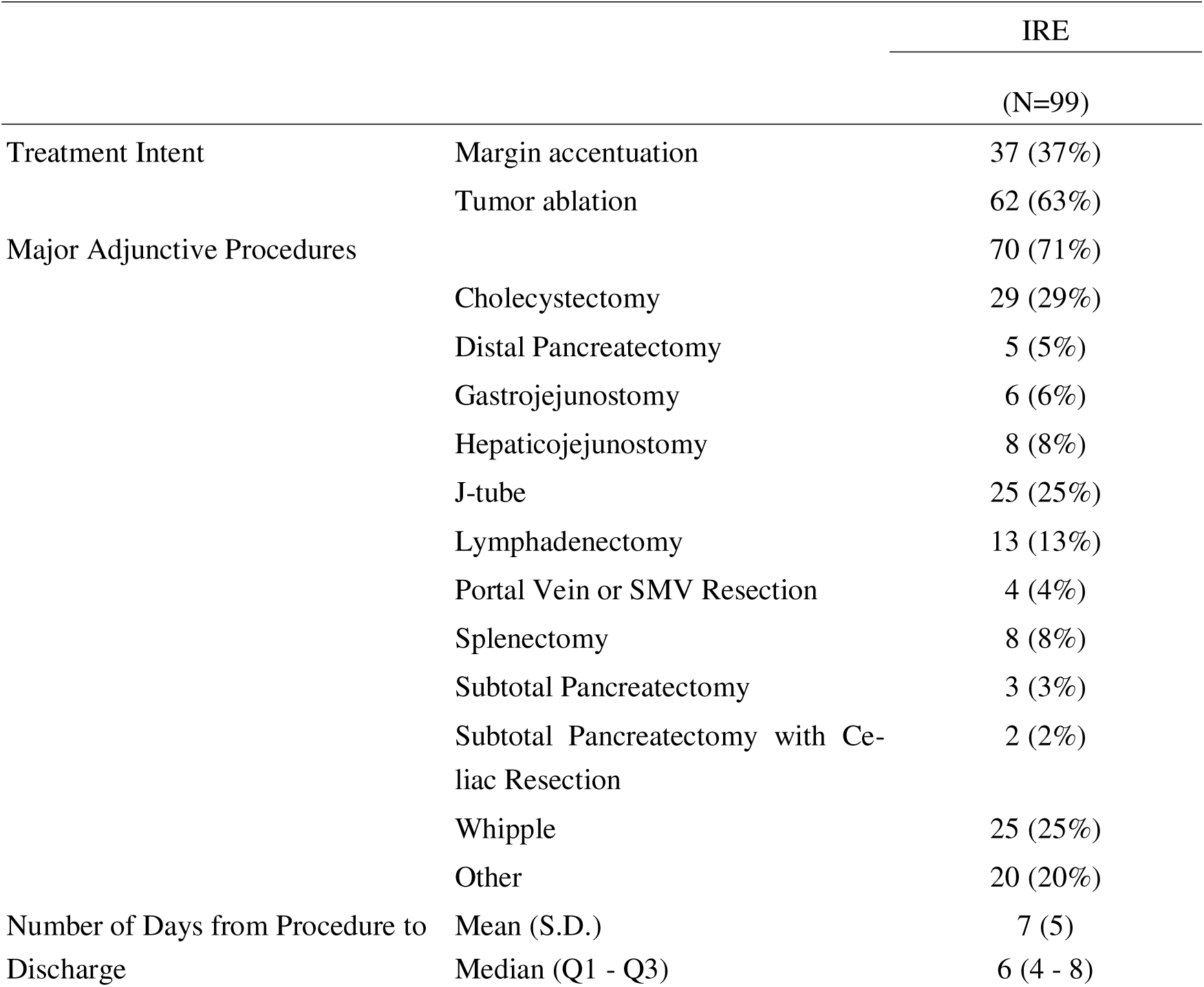
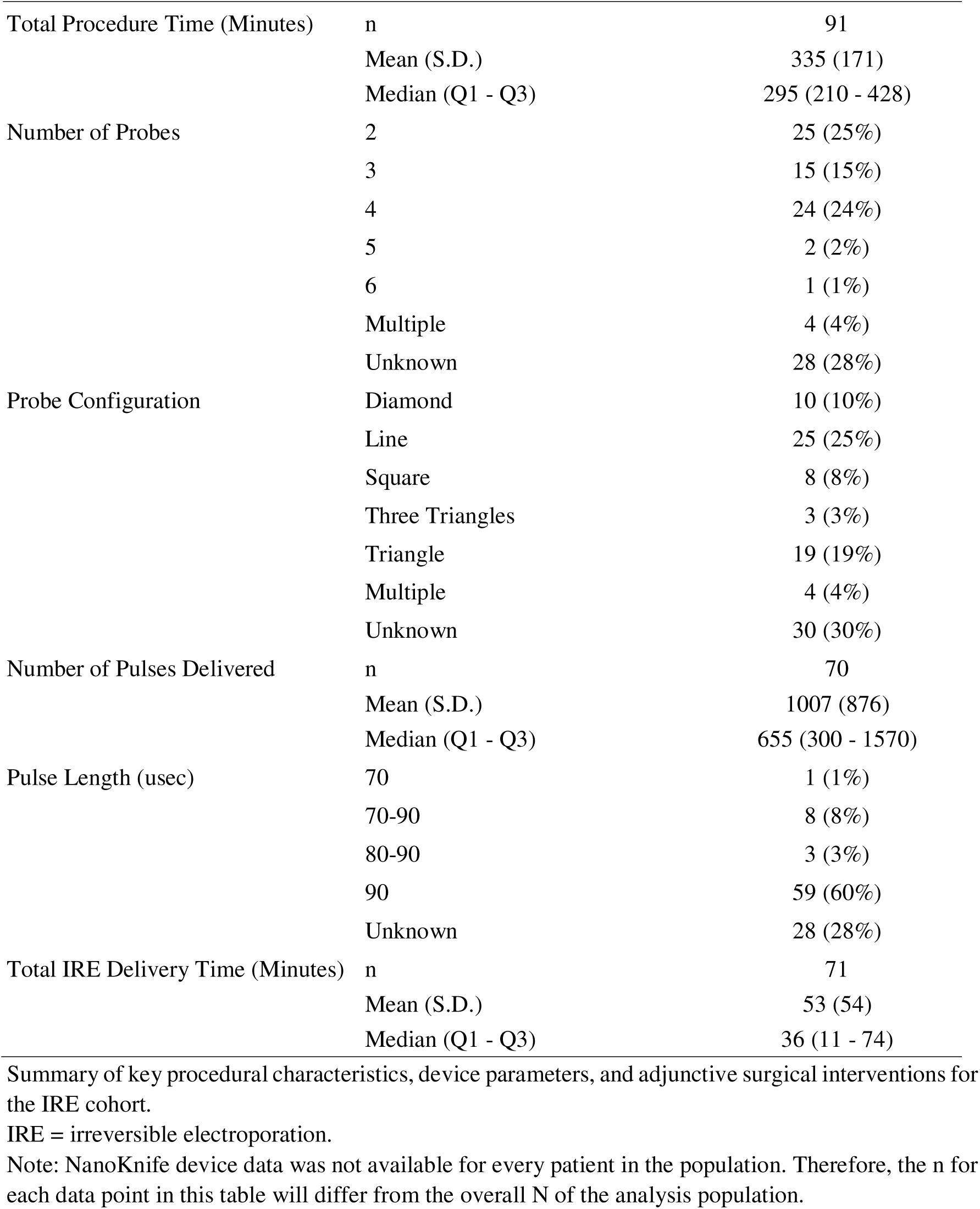
Procedure details for the IRE cohort.

### Post-Enrollment Chemotherapy and Radiation therapy

There were similar dose density and duration of chemotherapy in both cohorts post-enrollment; 36% (36/99) patients in the IRE cohort and 74% (28/38) in the SOC cohort received a second line of chemotherapy with a similar median (IQR) number of cycles of chemotherapy between cohorts with 4 (3-7) for IRE cohort and 5 (4-8) for the SOC cohort (p=0.29). Type of chemotherapy received was similar between cohorts with most patients receiving gemcitabine/nab-paclitaxel (41%, 26/137) or FOLFIRINOX (36%, 23/137). Overall, more SOC patients received third through-fifth lines of chemotherapy. Among patients with a known number of chemotherapy cycles, the median (IQR) cumulative number of cycles (first through third lines) was 6 (2-12) for the IRE cohort (n=85) and 11

(7-13) for the SOC cohort (n=37) (p=.01). Post-enrollment, 29% (29/99) of the IRE cohort received radiation and 40% (15/38) of the SOC cohort received radiation (p=0.02). Combining pre- and post-enrollment therapy, 65% (64/99) of IRE patients received radiation and 42% (16/38) of SOC patients received radiation (p=0.004).

### Overall Survival and Disease-Free Survival

The median OS from enrollment was 18 months (95% CI: 15-24) for IRE and 10 months (95% CI: 8-14) for the SOC cohort (p<0.0001) (Fig 1). Within the IRE cohort, OS was additionally calculated separately for patients receiving in-situ ablation (n=62) and margin accentuation (n=37) with a median OS of 18 months for the in-situ group (95% CI: 11-26) and 20 months for the margin accentuation group (95% CI: 14-28).

**Fig 1.**
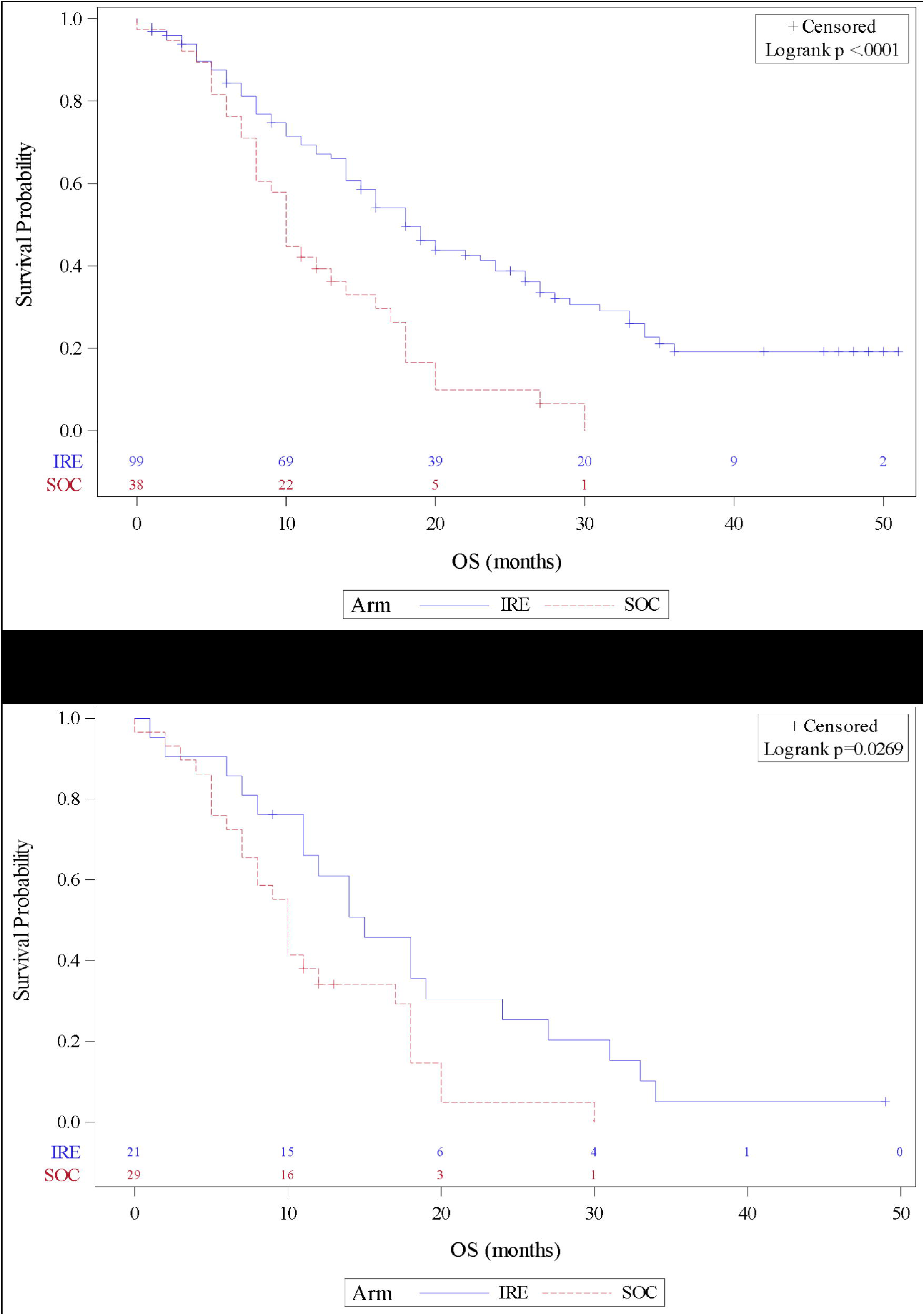
Product-limit survival estimates with number of patients at risk. The median overall survival from enrollment for irreversible electroporation (IRE) and for the standard of care (SOC) cohort. A) Overall survival by treatment arm. B) Overall survival by treatment arm, excluding patients with prolonged pre-enrollment.

In the sensitivity analysis reported in Table 3, the hazard ratio comparing OS of the IRE cohort to the SOC cohort was 0.4 (95% CI: 0.3-0.7; p<0.001). When calculating OS as time from diagnosis to death from any cause, the median OS in months was 28 (95% CI: 23-33) for the IRE cohort and 15 (95% CI: 12-21) for the SOC cohort (p<0.0001). Including only the prospectively enrolled SOC patients, the IRE and SOC cohorts had a median OS in months of 18 (95% CI:15-24) and 12 (95% CI: 0-27), re-Treatment received spectively (p=0.09). Excluding patients with prolonged pre-enrollment (greater than 6 months from diagnosis to enrollment), the median OS in months for IRE was 15 (95% CI: 11-24) and 10 (95% CI: 7-17) for the SOC cohort (p<0.05). Additional sensitivity analyses of OS and chemotherapy/radiation therapy can be found in S1 Table.

**Table 3.**
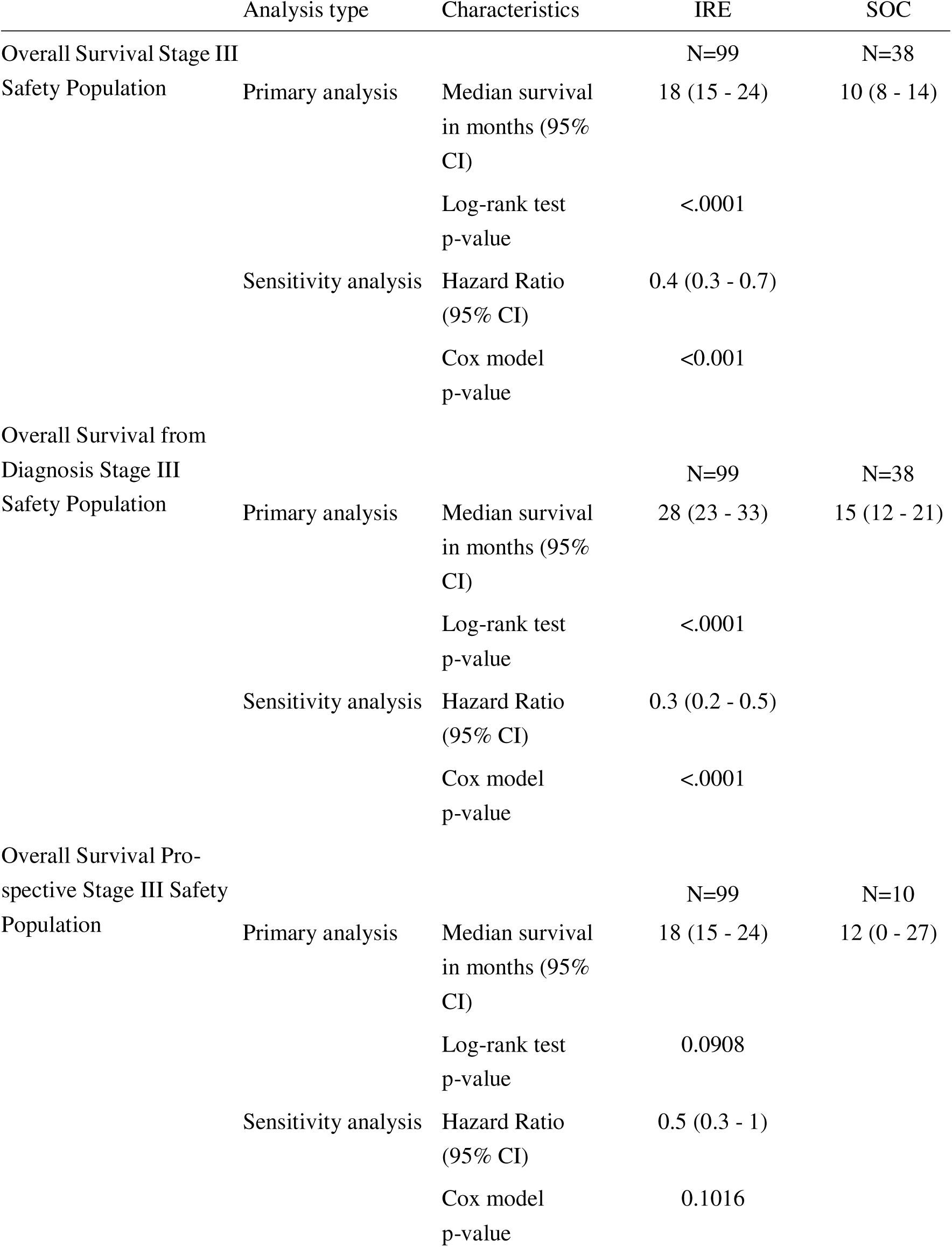
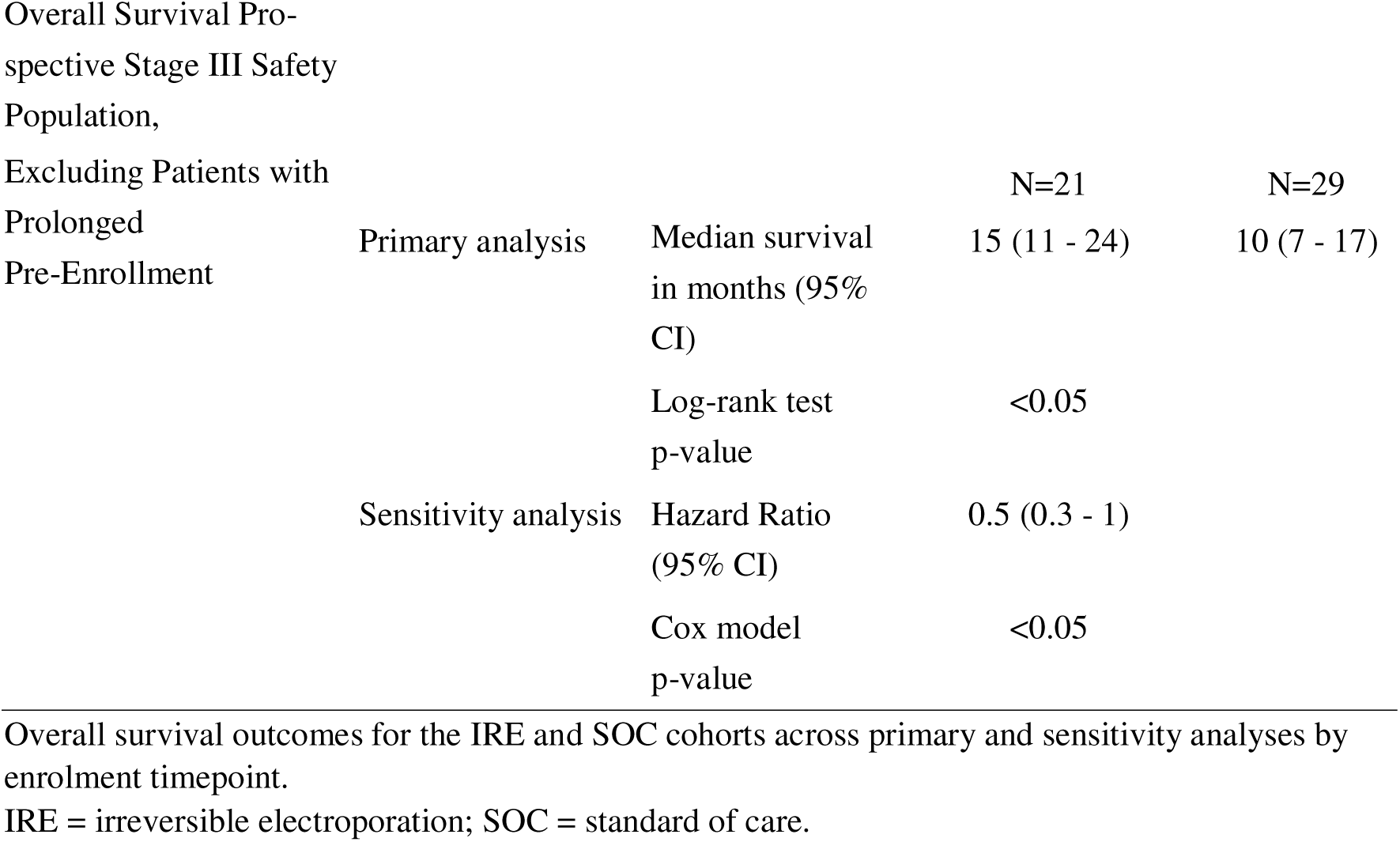
Overall survival, primary and sensitivity analyses.

The multivariate analysis of OS in Table 4 reported the hazard ratios for treatment (IRE versus SOC) as 0.3 (95% CI: 0.2-0.6; p<0.001), CA19-9 at baseline as 1 (95% CI: 1-1; p=0.58), anterior/posterior diameter as 1 (95% CI: 0.9-2; p=0.15), ECOG at baseline as 1 (95% CI: 0.7-2; p=0.73), and time from diagnosis to enrollment as 1 (95% CI: 1-1; p=0.14).

**Table 4.** Multivariate analysis of overall survival stage III safety population.

| Covariate | Characteristics |  |
| --- | --- | --- |
| Treatment | Hazard Ratio (95% CI) | 0.3 (0.2 - 0.6) |
|  | Cox model p-value | <0.001 |
| CA19-9 at Baseline | Hazard Ratio (95% CI) | 1 (1 - 1) |
|  | Cox model p-value | 0.58 |
| Anterior/Posterior Diameter (cm) | Hazard Ratio (95% CI) | 1 (0.9 - 2) |
|  | Cox model p-value | 0.15 |
| ECOG at Baseline | Hazard Ratio (95% CI) | 1 (0.7 - 2) |
|  | Cox model p-value | 0.73 |
| Time from Diagnosis to Enrollment | Hazard Ratio (95% CI) | 1 (1 - 1) |
|  | Cox model p-value | 0.14 |

Of the 12 IRE patients who were treated percutaneously, four patients had expired at the time of this analysis, with a mean survival of 12 months. Eight percutaneous patients were still alive, with a mean follow-up of 25 months.

Median PFS reported in Table 5, in months, was 9 (95% CI: 7-12) for the IRE cohort overall, and 6 (95% CI: 5-8) for SOC. The hazard ratio comparing PFS of the IRE cohort to the SOC cohort was 0.6 (95% CI: 0.4-0.9; p<0.05).

**Table 5.** Progression-free survival stage III safety population.

| Analysis type | Characteristics | Treatment received |  |
| --- | --- | --- | --- |
|  |  | IRE<br>(N=99) | SOC<br>(N=38) |
| Primary analysis | Median survival in months<br>(95% CI) | 9 (7 - 12) | 6 (5 - 8) |
|  | Log-rank test p-value | <0.01 |  |
| Sensitivity analysis | Hazard Ratio (95% CI) | 0.6 (0.4 - 1) |  |
|  | Cox model p-value | <0.05 |  |
| Disease Status | Censored at Date of Last<br>Follow-Up | 14 (14%) | 2 (5%) |
|  | Death | 27 (27%) | 10 (26%) |
|  | Disease Progression | 58 (59%) | 26 (68%) |
| Type of Progression | Distant | 39 (67%) | 12 (46%) |
|  | Local | 15 (26%) | 12 (46%) |
|  | Regional | 2 (3%) | 2 (8%) |
|  | Unknown | 2 (3%) | 0 (0%) |
| Location of Metastases<sup>a</sup> | Bone | 1 (3%) | 1 (8%) |
|  | Liver | 15 (39%) | 6 (50%) |
|  | Lung | 6 (15%) | 2 (17%) |
|  | Peritoneum | 11 (28%) | 5 (42%) |
|  | Other | 6 (15%) | 1 (8%) |

### Safety

Overall, 80% (79/99) of IRE patients and 95% (36/38) of SOC patients experienced an AE after enrollment. The timing of AEs from enrollment is shown in Table 6. For the IRE cohort, 29% (29/99) of patients had IRE-related AE with abdominal pain (8%, 8/99), fatigue (5%, 5/99), nausea (3% 3/99), pancreatic fistula (3%, 3/99), and vomiting (3%, 3/99) being the most common. A larger proportion of the SOC cohort had chemotherapy-related AEs than the IRE cohort: 68% (26/38) vs. 22% (22/99).

**Table 6.** Adverse events by enrollment timepoint.

| Timepoint | Treatment received<br>Number (%) of Patients |  | P-Value |
| --- | --- | --- | --- |
|  | IRE<br>(N=99) | SOC<br>(N=38) |  |
| Overall | 79 (80%) | 36 (95%) | 0.03 |
| 0-7 Days | 27 (27%) | 9 (24%) |  |
| 8-30 Days | 48 (49%) | 19 (50%) |  |
| 31-90 Days | 46 (47%) | 26 (68%) |  |
| 91-180 Days | 34 (34%) | 19 (50%) |  |
| >180 Days | 44 (44%) | 22 (58%) | 0.7 |

Total, 56% (55/99) of the IRE cohort and 71% (27/38) of the SOC cohort had a Grade 3 or greater AE. The most common Grade 3 or greater AEs for the IRE cohort were anemia (11%, 11/99), sepsis (8%, 8/99), and gastrointestinal hemorrhage (7%, 7/99), and for the SOC cohort were abdominal pain (13%, 5/38), ascites (13%, 5/38), anemia (11%, 4/38), sepsis (11%, 4/38), hypokalaemia (11%, 4/38), and diarrhea (11%, 4/38) (Table 7).

**Table 7.**
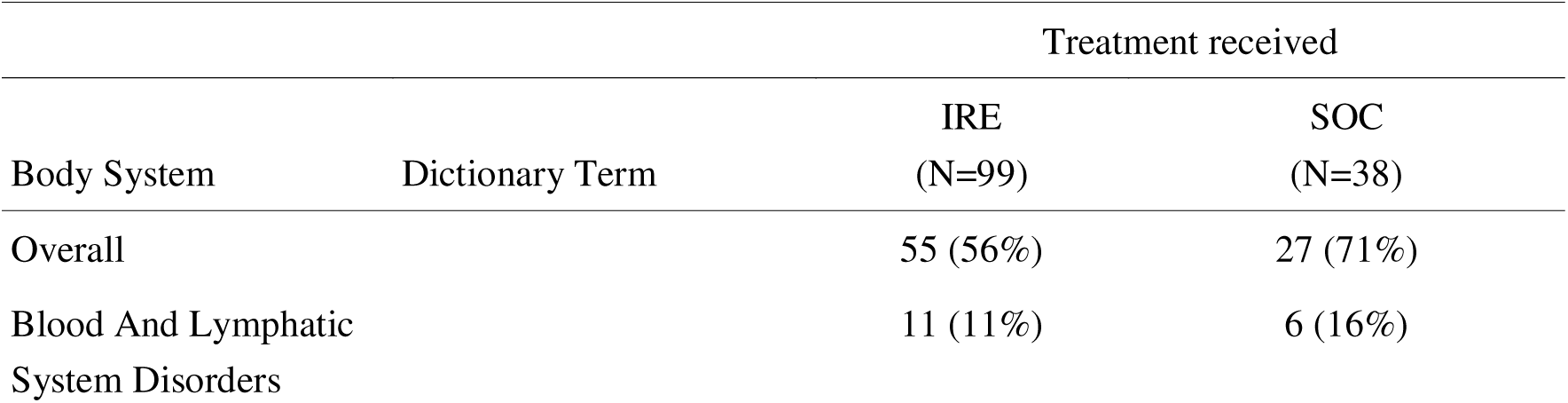
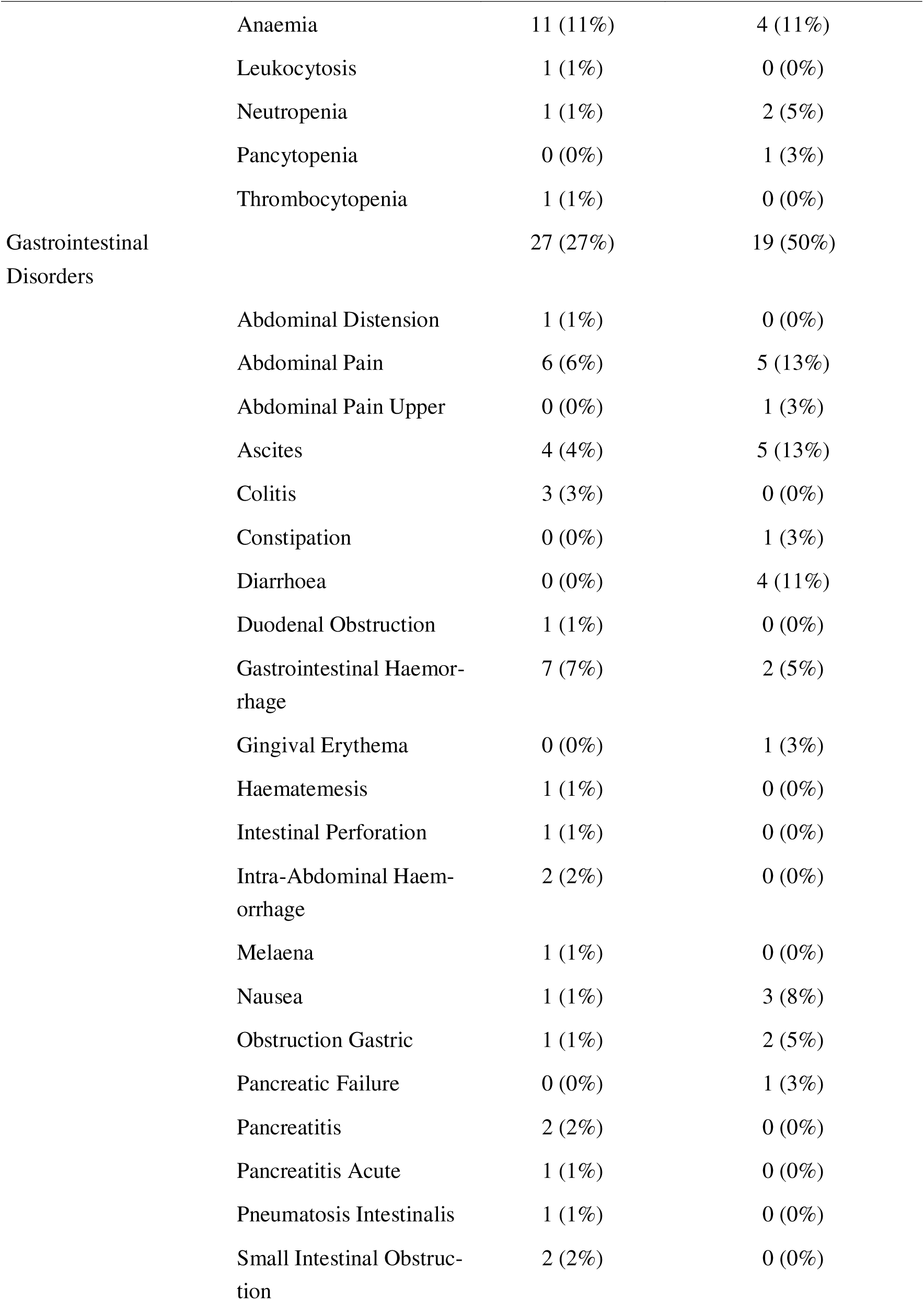
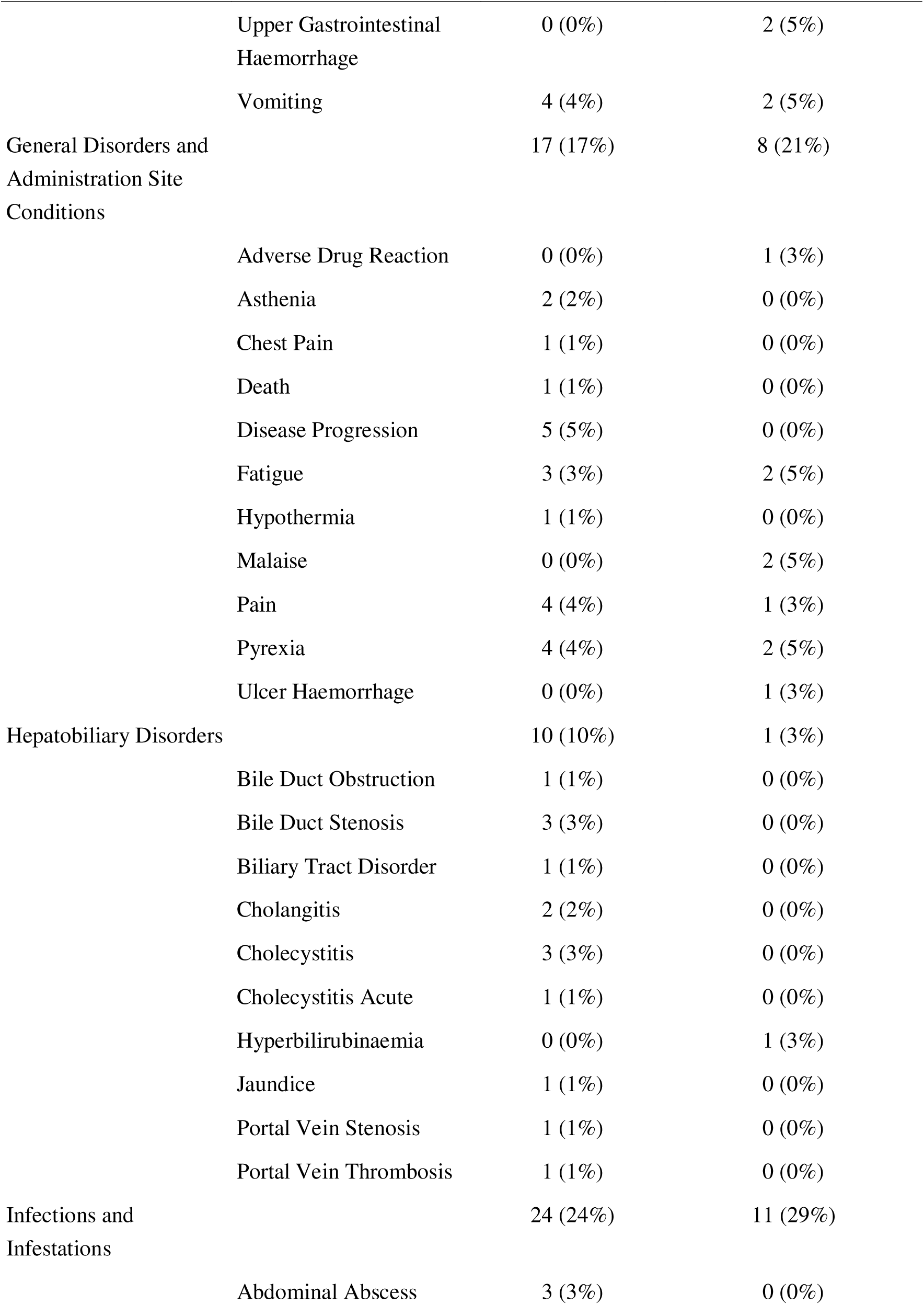
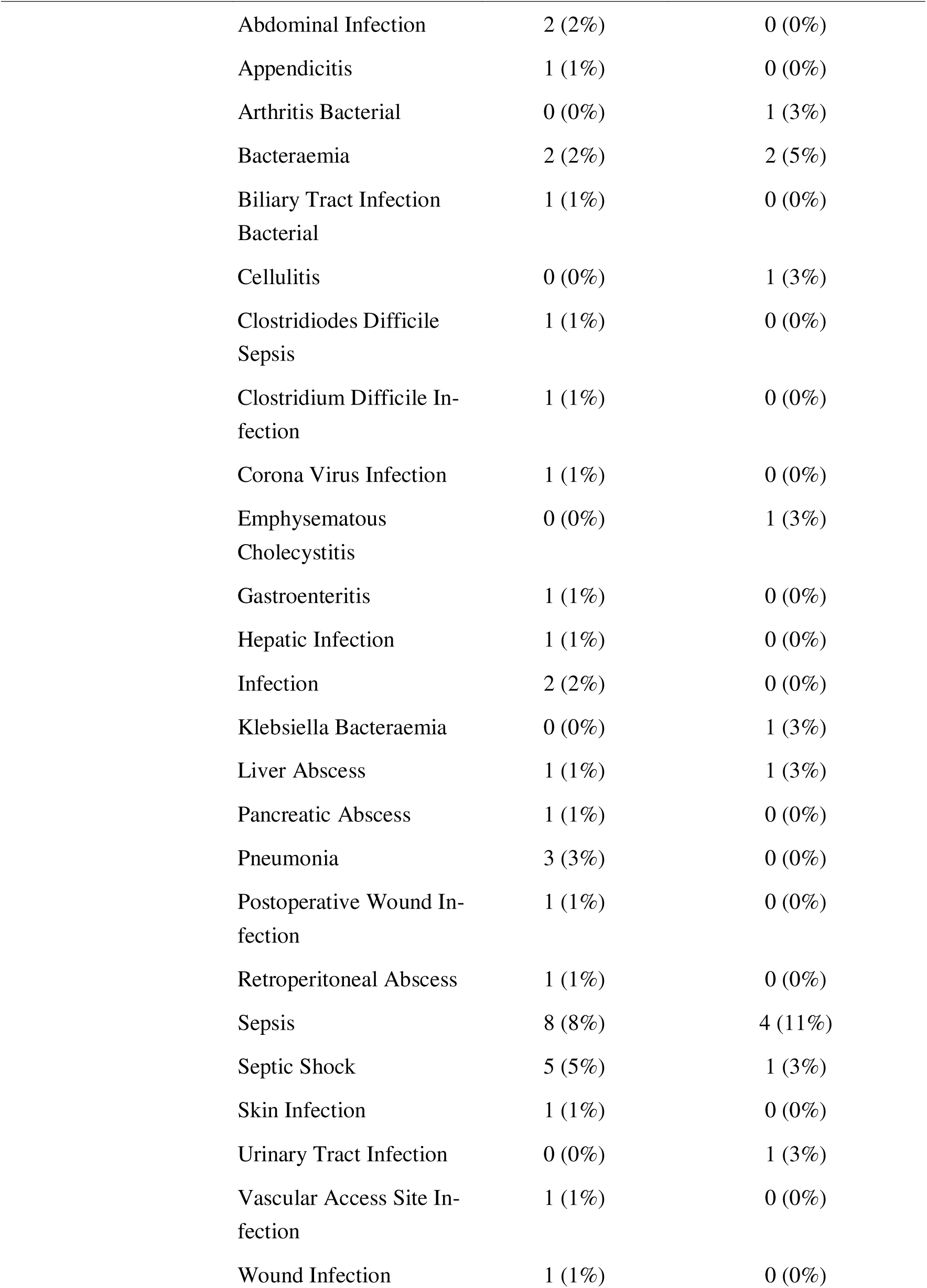
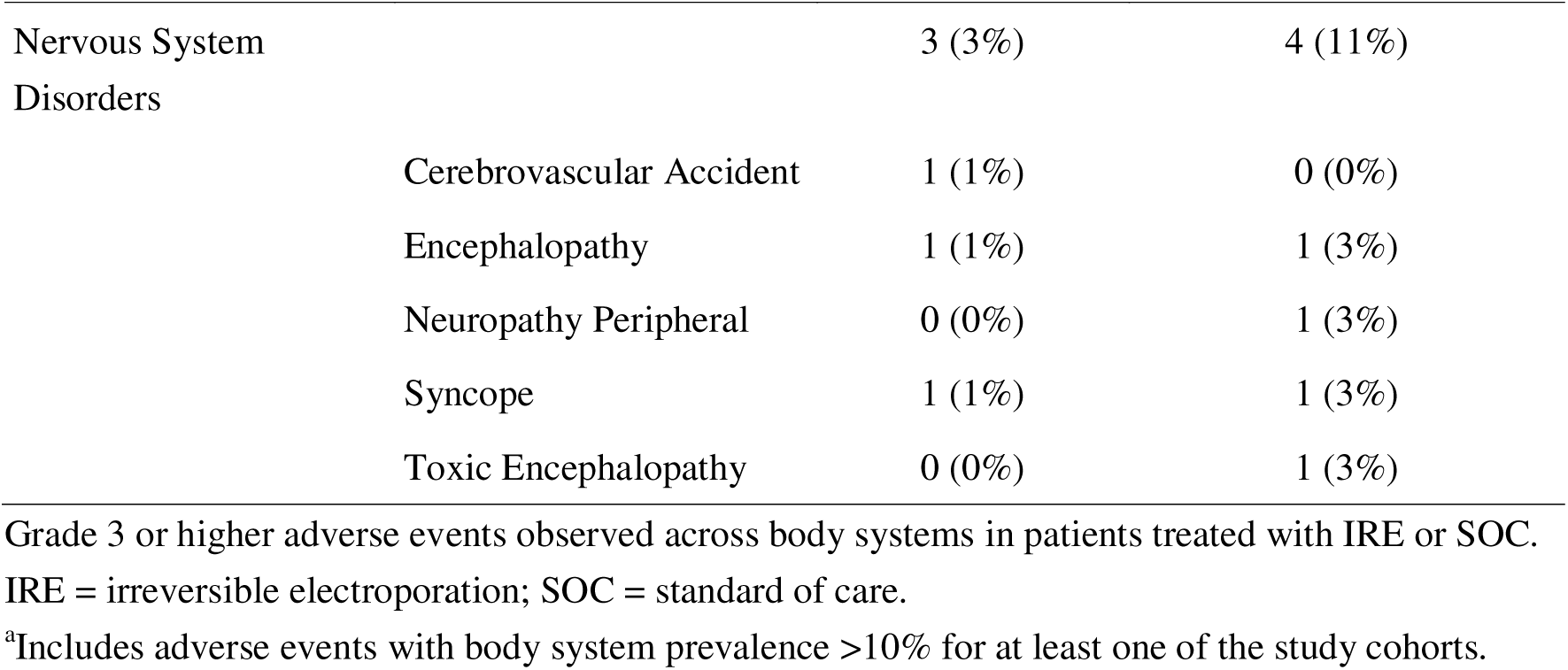
Grade 3+ adverse events experienced by the patient population.

## Discussion

Pancreatic cancer typically has no symptoms until reaching advanced stages and is often diagnosed when it is no longer resectable, leaving patients with chemotherapy and/or radiation treatment options and poor prognosis [1–3}. IRE presents an adjunctive option; DIRECT’s large registry shows IRE improved OS compared to SOC, halving the risk of death.

A prospective, registry study of 200 patients with locally advanced pancreatic cancer (LAPC) with 150 patients undergoing IRE alone and 50 with pancreatic resection plus IRE for margin accentuation using the open approach reported a median OS from diagnosis of 25 months [16]. The American Hepato-Pancreato-Biliary Association (AHPBA) Pancreatic Registry study, a prospective, multinational study of 152 patients with LAPC treated with IRE (alone or with resection) using the open approach reported a similar mean OS from diagnosis of 31 months [20]. Results from both studies are similar to the OS from diagnosis reported in the current study (28 months). While neither of these studies included a comparator cohort, results from both are more favorable than the median OS of patients with unresectable advanced pancreatic cancer receiving FOLFIRNOX (target median OS of 15 months from PANFIRE-2 trial [17]) suggesting a survival benefit over SOC. While non-comparative studies often report OS from diagnosis, for the primary analysis in this study which compared two treatment groups, survival time began at enrollment after patients in both cohorts received three months of SOC rather than time of diagnosis. Notably, the IRE cohort had a longer interval of nine months from diagnosis to enrollment, compared to only three months for the SOC cohort. This raises the potential for two forms of bias, lead-time bias that would favor the SOC group and immortal time bias that would favor the IRE group. In Stage III pancreatic cancer, patients who remain stable enough to be considered for IRE generally reflect a biologically distinct subset with controlled disease on induction therapy.

This has never been reported in Stage III PDA and this yes may lead to some bias, it also further emphasizes the need to consider effective local consolidative therapies in this stage disease. Our analyses aim to separate this biology-driven selection effect from a procedure-related effect; however, we acknowledge this cannot be fully eliminated in a non-randomized design.

Results from the current study are consistent with comparative studies of IRE to other treatment approaches. A single-center study used propensity score matching to compare 36 matched pairs of patients with LAPC treated with open IRE or radiotherapy after induction chemotherapy. In this study, the IRE group had a median OS of 22 months compared to 10.6 months for the radiotherapy group.

Median PFS was 8 months for the IRE group which is a slightly shorter duration than the median PFS observed in the current study (10 months) [21]. The CROSSFIRE trial, a single-center, open-label, randomized controlled trial (RCT) comparing MRI-guided stereotactic ablative body radiotherapy (SABR) to CT-guided percutaneous IRE in 68 patients with locally advanced pancreatic cancer reported a median OS from randomization of 16 months in the SABR group compared to 13 months in the IRE group though this difference was not statistically significant. Furthermore, there were no detected differences in severe AEs between the treatment groups [22].

In the current study, a lower proportion of patients in the IRE cohort had AEs overall compared to the SOC cohort (80%, 79/99 vs. 95%, 36/38). The IRE cohort also had a lower incidence of AEs that were Grade 3+ (56%, 55/99 vs. 71%, 27/38) suggesting that IRE has a superior safety profile over SOC. However, the percentage of patients with a Grade 3+ AE was higher than that of studies reported in a systematic review of IRE which ranged from 1% to 42%. While this range is considerable, the reasons for this variation may be due to differences in treatment protocols, size of treated tumors, physician experience with the IRE procedure, or method of delivery (open vs. percutaneous) [23]. Additionally, this prospective IDE was rigorously designed to collect AE data and therefore may have been more sensitive to the capture of AEs compared to smaller retrospective studies. Regarding the high rate of AEs in the SOC cohort, patients with pancreatic cancer tend to experience AEs due to the severity of their illness especially as the disease progresses post-treatment. As an example, the percentage of patients with a Grade 3+ AE in the SOC cohort (71%, 27/38) was comparable to the rate of grade 3 or 4 AEs among patients randomized to modified FOLFIRINOX (76%) in an RCT randomizing patients to adjuvant chemotherapy [24].

The DIRECT Registry study was not designed to compare IRE with radiation or preoperative radiation; rather, it was designed to highlight a potential role for IRE as an adjunct or alternative in selected cases, as optimal local therapy for patients with unresectable disease or margins-at-risk remains an unresolved question. Future randomized studies and multidisciplinary decision-making would be essential to define the optional sequencing and selection of local therapies.

We acknowledge the imbalance in sample size between the SOC and IRE cohorts. This reflects real-world treatment allocation and differential eligibility for IRE rather than selective enrollment. Importantly, unequal group sizes do not introduce bias per se but may affect statistical power and precision of effect estimates. To address this, we used appropriate statistical methods that do not assume equal sample sizes and performed adjusted analyses to account for baseline differences. Sensitivity analyses demonstrated consistent results, supporting the robustness of our findings despite sample size imbalance.

This study had several limitations. The study experienced difficulty with enrollment into the SOC cohort as some patients specifically sought IRE treatment. There is also heterogeneity within the SOC cohort as there is no standardized SOC for Stage 3 PDAC after induction chemotherapy. While results show that IRE performed better than SOC in terms of survival and safety, the treatments received by patients in the SOC cohort may vary. Chemotherapy received post-enrollment was similar between the IRE and SOC cohorts, though the SOC cohort received more mean cumulative cycles of chemotherapy. Additionally, while IRE patients were more likely to receive radiation pre-enrollment, and SOC patients were more likely to receive radiation post-enrollment, a larger proportion of patients in the IRE cohort received radiation overall. This study also was not able to fully exclude residual immortal time bias. Finally, as previously mentioned, there is potential for lead-time bias favoring the SOC group and immortal time bias favoring the IRE group.

### Conclusion

In the DIRECT Registry, IRE resulted in improved OS and a more favorable safety profile than SOC in patients with Stage 3 PDAC demonstrating that IRE is a safe and effective treatment option for patients with unresectable disease. Future research may compare IRE to other treatment modalities as well as assess differences between the open and percutaneous approaches.

## Data Availability

All data produced in the present work are contained in the manuscript

## Supporting information

**S1 Fig. Attrition diagram.** There was no attrition as all 137 patients who were enrolled completed the study.

**S2 Fig. Study design scheme.** The outline of the clinical pathway for patients diagnosed with Stage 3 Pancreatic Ductal Adenocarcinoma.

**S1 Table. Sensitivity analysis of overall survival.** Sensitivity analyses evaluating overall survival according to chemotherapy and radiation therapy.

**S1 Methods. Study eligibility criteria for the DIRECT Registry.** Patient inclusion and exclusion criteria.

## Notes

### Competing Interest Statement

Dr. Martin has received consulting fees and reimbursement for travel expenses from
AngioDynamics. Dr. Narayanan received consulting fees for AngioDynamics, Stryker
and Varian Inter-ventional Solutions and is part of the advisory board of Quantum
Surgical and Betaglue. Dr. Kluger has received consulting fees from Histosonics. Dr.
Iannatti has received consulting fees from Ethicon, Medtronic, BK Medical, Dilon,
Boston Scientific, and AngioDynamics. Dr. Heithaus is on a speaker bureau for
AngioDynamics, Boston Scientific, Canon Medical, and Penumbra and has received
research funding from Penumbra, Canon Medical, and AngioDynamics. Dr. Polanco
has received honoraria from Intuitive Surgical and consulting fees from Iota
Biomedical. Dr. White reports receiving research funding from Trisalus Life Sciences.
All other authors report having no interests to declare.

### Clinical Trial

NCT03899649

### Funding Statement

The study sponsor (AngioDynamics, Inc.; https://www.angiodynamics.com) provided
financial and logistical support but had no role in data collection or data interpretation.
The academic investigators (RCGM, RRW, MMB, GN, MDK, DAI, PMP, CWH, SPC,
REH, TW, CHFC) had full access to the complete dataset and retained final
responsibility for data interpretation and the decision to submit the manuscript for
publication.

### Author Declarations

The study received approval from the Western Institutional Review Board (approval 2019- 140 0965). Institutional Review Board of the University of Louisville, Northwest Community Healthcare, Illinois Center for Pancreatic and Hepatobiliary Diseases, USF Health South Tampa Center for Advanced Healthcare, USF Health Morsani Center for Advanced Healthcare, Baptist Hospital of Miami, Miami Cancer Institute, University of California San Diego, Atrium/Carolinas Healthcare System, Columbia University, UT Southwestern Medical Center, Barnes-Jewish Hospital, Washington University School of Medicine, Center for Advanced Medicine, Mayo Clinic, WellStar Summit Surgical, Capital Health System, St. Lukes University Health Network, University of Iowa, Augusta University, University of Florida, and NYU Langone Health gave ethical approval for this work.

