## Supplemental Figure 1 for "Irreversible electroporation associated with improved overall survival vs standard of care for stage 3 pancreatic ductal adenocarcinoma"

**S1 Fig. Attrition diagram.** There was no attrition as all 137 patients who were enrolled completed the study. IRE = irreversible electroporation; SOC = standard of care.

### Enrollment

Analyzed (n=99)
 Excluded from analysis (n=0)

Lost to follow-up (n=0)

Discontinued IRE (n=0)

Enrolled (n=137)

Assessed for eligibility (n=137)

Excluded (n=0)

  Not meeting inclusion criteria (n=0)

  Declined to participate (n=0)

  Other reasons (n=0)

### Allocation

Allocated to IRE (n=99)

 Received IRE (n=99)

 Did not receive allocated intervention (n=0)

Allocated to SOC (n=38)

 Prospectively allocated (n=10)

 Retrospectively allocated (n=28)

Site 001 (n=17)

Site 068 (n=11)

### Follow-Up

Lost to follow-up (n=0)

Discontinued intervention (n=0)

### Analysis

Analyzed (n=38)
 Excluded from analysis (n=0)
