## Supplemental Methods for "Irreversible electroporation associated with improved overall survival vs standard of care for stage 3 pancreatic ductal adenocarcinoma"

**Supplementary Methods. Study eligibility criteria for the DIRECT Registry.** Patient inclusion and exclusion criteria.

**Inclusion Criteria**

All the following criteria were met in order to be enrolled and continue participation in the study:

1. Provision of signed and dated informed consent form

2. Patient was 18 years of age and older

3. Patient had a diagnosis of Stage 3 pancreatic (PC) cytologically or pathologically confirmed per American Joint Committee on Cancer (AJCC) staging criteria

4. Patient had a tumor evaluated as Stage 3 according to National Comprehensive Cancer Network (NCCN) guidelines, based on radiographic imaging or exploratory surgery

5. Maximum axial and anterior to posterior tumor dimension of ≤3.5 cm after standard of care (SOC)

6. Patient received three months of SOC per each participating institution’s guidelines

7. Patient had an Eastern Cooperative Oncology Group (ECOG) performance status of 0 or 1

8. Patient had an American Society of Anesthesiologists (ASA) classification of physical health

status of 1, 2, 3 or 4

9. Patients at IRE sites who were deemed eligible for IRE received ablation using the NanoKnife System

10. Patient showed no evidence of disease progression based on NCCN guidelines after completing three months of SOC

**Exclusion Criteria**

A potential patient was excluded from the study if he/she met any of the following exclusion criteria:

1. Participation in an interventional trial for PC during the study data collection period

2. Pregnant or lactating patients or male or female patients of reproductive potential who were not willing to employ highly effective birth control from screening to 6 months after the last dose of chemotherapy

3. Patients who were unable to tolerate general anesthetic with full skeletal muscle blockade

4. Patients with the presence of implanted cardiac pacemakers, defibrillators, electronic devices or implanted devices with metal parts in the thoracic cavity at the time of IRE
