## Supplemental Table 1 for "Irreversible electroporation associated with improved overall survival vs standard of care for stage 3 pancreatic ductal adenocarcinoma"

**S1 Table. Sensitivity analysis of overall survival.**

| Model | Covariate | Characteristics |  |
| --- | --- | --- | --- |
| Overall Survival and Chemotherapy | Treatment | Hazard Ratio (95% CI) | 0.3 (0.2 - 0.6) |
|  |  | Cox model p-value | <.0001 |
|  | Cumulative Number of Chemotherapy Cycles | Hazard Ratio (95% CI) | 1 (0.9 - 1) |
|  |  | Cox model p-value | <0.05 |
| Overall Survival and Radiation Therapy | Treatment | Hazard Ratio (95% CI) | 0.5 (0.3 - 0.8) |
|  |  | Cox model p-value | <0.005 |
|  | Radiation Therapy, Yes vs No | Hazard Ratio (95% CI) | 2 (1 - 2) |
|  |  | Cox model p-value | 0.05 |
|  | Radiation Therapy, Yes vs Unknown | Hazard Ratio (95% CI) | 3 (2 - 6) |
|  |  | Cox model p-value | <0.01 |

Sensitivity analyses evaluating overall survival according to chemotherapy and radiation therapy.
