## Supplemental Figure 2 for "Irreversible electroporation associated with improved overall survival vs standard of care for stage 3 pancreatic ductal adenocarcinoma"

**S2 Fig. Study design scheme**. The outline of the clinical pathway for patients diagnosed with Stage 3 Pancreatic Ductal Adenocarcinoma. AE = adverse event; IRE = irreversible electroporation.

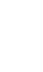

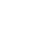

Diagnosis of Stage 3 Pancreatic Ductal Adenocarcinoma

3-Months of Standard of Care

Informed Consent and Baseline Data Collection

Non-IRE

IRE

Monthly and Quarterly Data Collection

Death, Discontinuation, Loss to Follow-up, Study End

AE
